# Mindfulness related changes in grey matter: A Systematic Review and Meta-analysis

**DOI:** 10.1101/2020.03.31.20049510

**Authors:** Cyril R. Pernet, Nikolai Belov, Arnaud Delorme, Alison Zammit

## Abstract

Knowing target regions undergoing structural changes caused by behavioural interventions is paramount in evaluating the effectiveness of such practices. Here, using a systematic review approach, we identified 25 peer-reviewed magnetic resonance imaging (MRI) studies demonstrating grey matter changes related to mindfulness meditation. An activation likelihood estimation (ALE) analysis (n=16) revealed the right anterior ventral insula as the only significant region with consistent effect across studies, whilst an additional functional connectivity analysis indicates that both left and right insulae, and the anterior cingulate gyrus with adjacent paracingulate gyri should also be considered in future studies. Statistical meta-analyses suggest medium to strong effect sizes from Cohen’s d ∼0.8 in the right insula to ∼1 using maxima across the whole brain. The systematic review revealed design issues with selection, information, attrition and confirmation biases, in addition to weak statistical power. In conclusion, our analyses show that mindfulness meditation practice does induce grey matter changes but also that improvements in methodology are needed to establish mindfulness as a therapeutic intervention.

## Introduction

The relatively recent increase in the popularity of meditation in the Western world has received much attention, with many claiming associated mental health benefits. Concerns about the qualities of studies and potential harm to patients have nevertheless also been raised (Farias & Wikholm, 2016) given that up to a quarter of meditators have reported unpleasant and potentially deleterious meditation-related experiences (Schlosser et al., 2019). Medical intervention using meditation (in various forms) has been used predominantly in the treatment of chronic pain, and to a lesser extent in the treatment of obesity, aging, dementia, and psychiatric disorders. Literature reviews and meta analyses show some evidence for lowering perceived pain along with improving associated symptoms such as depression and anxiety (Edwards & Loprinzi, 2018) or sleep disorders (Kwekkeboom & Bratzke, 2016), usually with no change in actual symptoms (Hood & Jedel, 2017; see however Ball, Nur Shafina Muhammad Sharizan, Franklin, & Rogozinska, 2017). Such intervention seems also ineffective in ‘heavy’ patients such as those in palliative care (Latorraca et al., 2017). Meditation has shown benefits in promoting better eating behaviours therefore assisting weight regulation (Dunn et al., 2018), enhancing cognitive efficiency in healthy aging (Sperduti et al., 2017) and reducing cognitive decline in dementia (Russell-Williams et al., 2018). Finally, there is also evidence that meditation improves patients with mental illnesses, with reductions in psychotic symptoms, depression scores, and improved level of functioning (Potes et al., 2018). In most reviews, meditation has been shown to improve quality of life (Edwards & Loprinzi, 2018; Hilton et al., 2017; Hood & Jedel, 2017; Potes et al., 2018; Russell-Williams et al., 2018). Accumulating evidence for benefits, risks and mechanisms of meditation is therefore valuable to improve patients’ health whilst reducing interventions and healthcare costs (Innes & Selfe, 2014).

ElectroEncephaloGraphy (EEG) and structural/functional MRI have been used extensively to study brain changes underpinning behavioural effects of meditation. A previous meta-analysis by Boccia et al. (2015) suggests functional changes in meditators relative to controls in the thalamus, striatum and frontal cortex along with structural changes in some of these regions. The mechanism leading to grey matter (GM) changes is unknown, although EEG findings support the hypothesis that meditation increases neuronal communication which in turn causes GM changes via synaptic plasticity. For instance, meditation tends to increase fast oscillations of 12–30 Hz, which are usually associated with high vigilance levels (Braboszcz & Delorme, 2011; Laufs et al., 2006) and increase in phase-locking gamma-band oscillations (Lutz et al., 2004). Such increases in neural communication and connectivity might mediate observed long term brain structural changes.

Our review focused on studies looking at GM and aimed at (1) investigating the strength of reported associations and (2) establishing evidence for local GM changes due to meditation. Meditation is here strictly defined as the practice of focused attention or awareness with the aim of cultivating a calm and stable mental state, which typically corresponds to what has been termed mindfulness. Consequently, we excluded from our analyses any studies that included movement therapies, such as yoga alongside meditation (Boccia et al., 2015; Fox et al., 2014; Last et al., 2017) since these additional practices may act as confounding factors (Colcombe et al., 2006; Erickson et al., 2014). Since proposed meditation interventions often hypothesize brain changes that will regulate different aspects of behaviour, it is essential to establish which brain regions show changes. For example, if patients show an alteration in one brain region, imaging can address whether an intervention may re-establish a normal range of values in this region or if behavioural changes are mediated by compensatory mechanisms (Bishop, 2013). Although many brain changes are functional by nature (change in activation level, frequency of activity or activity coupling), we focused here on structural Grey Matter (GM) changes only, because it provides evidence of neural plasticity for long term mental health changes.

## Material and method

### Systematic Review

#### Search strategy

The Ovid interface was used to search MEDLINE, EMBASE, PsycINFO, AMED and Global Health. The following terms were searched separately on the 10th of October 2018, using the multi-field search tool with “All Fields” enabled: (1) Meditat* OR Mindfulness (2) Magnetic Resonance Imaging OR MRI OR MR Imaging OR sMRI OR Neuroimaging (3) Grey Matter OR Grey Matter OR GM OR Cortical Thickness OR Cortex OR Brain. Searches 1-3 were combined with a Boolean AND function. The high number of hits returned from PsycINFO was reduced by consulting relevant subject headings and performing an advanced search (as follows) with ‘map to subject headings’ and ‘Auto Explode’ enabled: (1) Meditation OR Mindfulness (2) Magnetic Resonance Imaging or Neuroimaging (3) Grey Matter or Cerebral Cortex. Again, searches 1-3 were combined using a Boolean AND function.

#### Inclusion Criteria

Articles included must have met the following criteria: (i) Involve meditation or mindfulness (ii) Use structural MRI (iii) Measure GM density, volume or thickness.

#### Exclusion criteria

To produce a focused report and avoid confounding effects, the exclusion criteria comprised of (i) Articles involving physical movement alongside meditation: For example, the mindfulness-based stress reduction (MBSR) program developed by Kabat-Zinn, which incorporates yoga (Santorelli et al., 2017), (ii) Articles where structural MRI was not performed, and (iii) Articles that do not measure grey matter volume (GMV)/density (GMD) or cortical thickness (CTh): Measurement outcomes such as WM changes and gyrification patterns were not included.

#### Bias Analysis

Articles were assessed for bias using the Clinical Appraisal Skills Programme checklists https://casp-uk.net/casp-tools-checklists/. We report here specifically on selection, information and attrition biases.

### Meta-Analyses

#### Amplitude of morphological changes?

For each study included, the maximum reported effect was extracted (e.g. t-values, regression slope or Pearsons’ r correlation coefficients) either directly from the text or inferred from figures. Importantly, the maximum effect size was used irrespective of the location in space, thus establishing a generic evidence for an effect of meditation on GM. For Chételat et al. (2017) and Murakami et al. (2012) data points were extracted from scatter plots using WebPlotDigitizer (Rohatgi, A., 2019) while for Kumar et al. (2014), t-value was taken from the maximum of the figure color scale. From these values and number of participants, Hedges’ g effect sizes were obtained and a random-effect meta-analysis (Maximum-likelihood as estimator) was computed using the *dmetar* R package (Harrer, M. et al., 2019). An additional meta-analysis was also conducted for the right insula, based on the a-posteriori observation of the spatial distribution of reported effects. From the meta-analyses effect sizes, Hedges’ g was transformed to Cohen’s d using equation 4 from Lakens (2013) and G-power (Faul et al., 2007, 2009) was used to estimate the number of subjects needed for 80% and 95% statistical power of a two-samples t-test.

#### Where in the brain can we see changes?

For each study showing structural brain differences in meditators, and when available, coordinates from significantly different regions were entered into an anatomic likelihood estimation analysis using GingerALE version 3.02 (Eickhoff et al., 2012; Eickhoff, S.B. et al., 2009; Turkeltaub et al., 2012), which estimates the likelihood that a region contains an effect taking into account both reported locations and sample sizes (here using the degrees of freedom of the test from which coordinates were used). Since the analysis is focused on spatial analyses, only whole brain studies are used, but using all coordinates. The ALE map was thresholded at a p-value of 0.05 after family-wise error cluster correction with 1000 permutations and the default p=0.001 cluster forming threshold. Since structure and function of the human brain are intricately linked through multiple levels and modes of brain connectivity (Sporn et al., 2005), we sought to gain further insight of meditation induced neuroplasticity by using functional connectivity maps from NeuroSynth (Yarkoni et al., 2011). The functional connectivity maps represent resting-state functional connectivity analysis performed on 1,000 human subjects (Buckner et al., 2014), with voxel seeds from 7 study coordinates in and around the region found significant by the ALE analysis. These coordinate-based connectivity maps were then binarized (thresholded at 0.2) and summed, and this final map thresholded for significance at 88%, i.e. above the 95% upper bound of the chance level confidence interval.

## Results

### Search Results

The total number of references gathered was 867 (MEDLINE=234, Embase=545 AMED= 3, Global Health= 2, PsycINFO= 83). After deleting duplicates this was reduced to 698. Of these, 530 were excluded following abstract screening based on the criteria defined previously. The full texts of 143 articles were examined, of which 25 met inclusion criteria (see table 1). Cross-checking with other reviews (e.g. (Boccia et al., 2015) revealed a high number of matches, which was taken as evidence of a good search strategy.

**Table 1.**
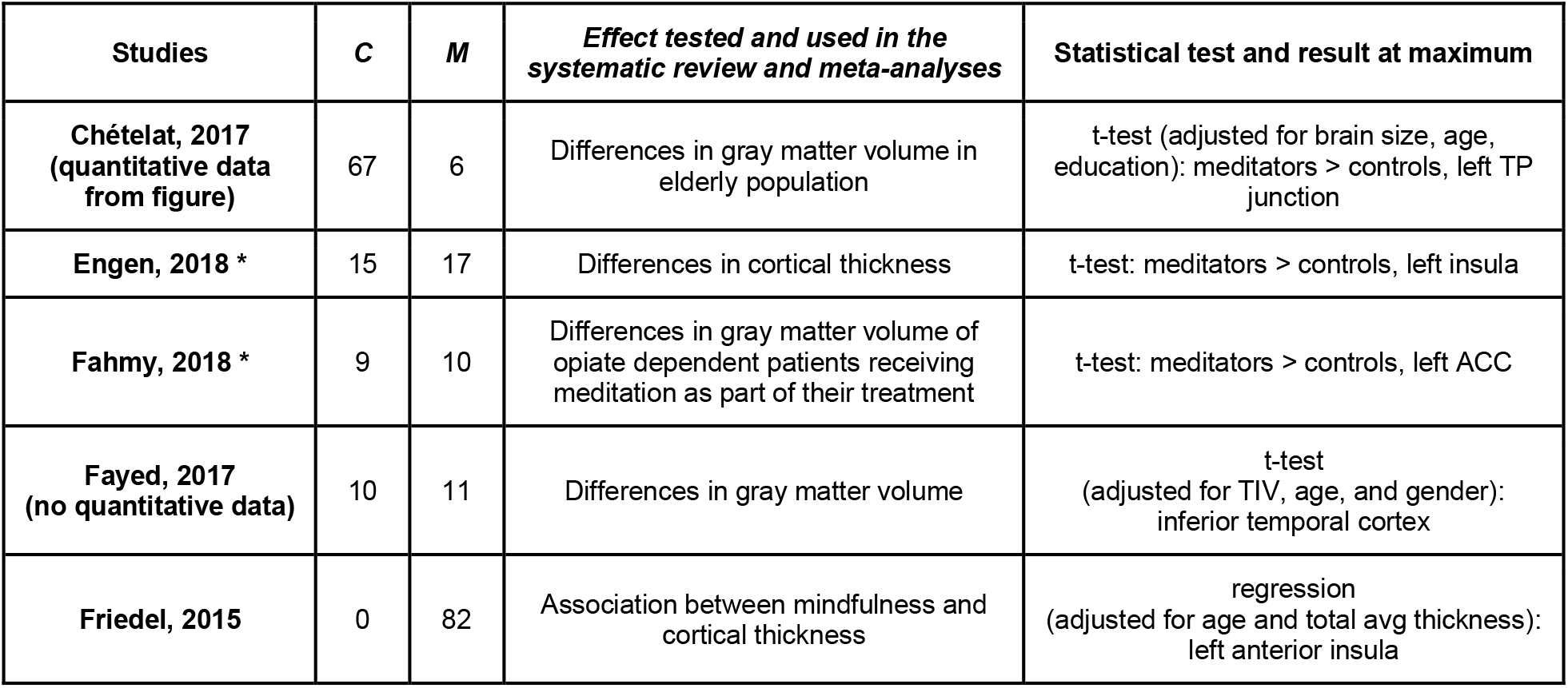
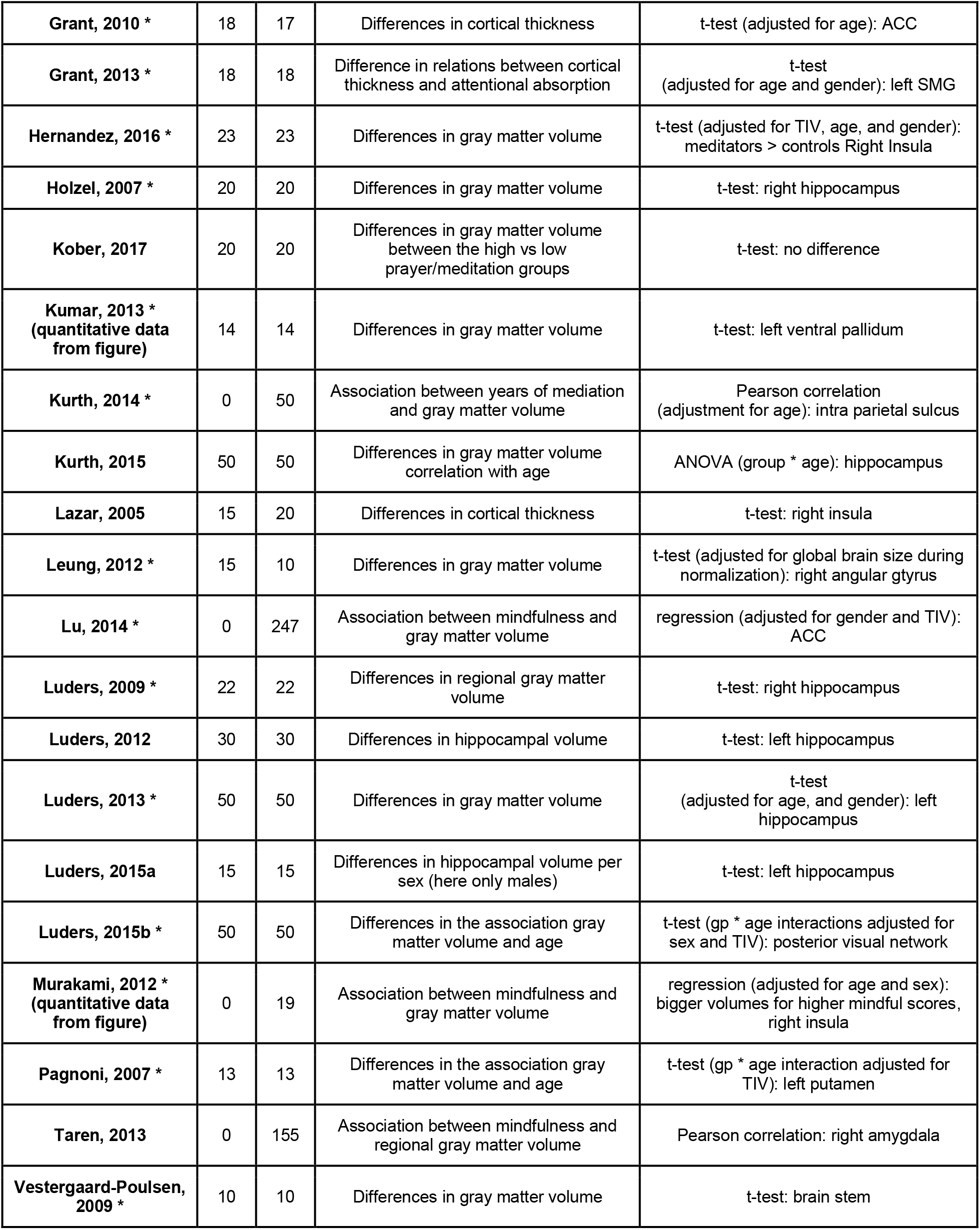
List of the 25 studies included in the systematic review (* denotes those included in the ALE analysis) with the number of control subjects (C), the number of meditators (M), and the effect tested and the corresponding statistical test and result (TP = temporo-parietal, ACC = Anterior Cingulate Cortex, SMG: supramarginal gyrus)

#### Study range and characteristics

The sum of the participants included in the 25 selected studies was 1646. Accounting for overlapping samples reduced this to 1406 participants (mean: 56.24, range: 19-247). Participant age ranged from 12 to 87 years old, and various meditation/mindfulness disciplines were studied.

### Strength of associations and bias

Twenty-three out the 25 included studies reported quantitative changes associated with meditation. Nineteen of them compared directly meditators to non-meditators (only 18 used in the analysis as 1 study only reported values of the difference between patients, even if controls were present) and the six remaining studies reporting associations with the amount of meditation. The meta-analysis random-effect-model across all 23 studies from which we had data, for the strongest effect independently of the brain area and design, showed an estimated effect of g = 0.8244 [95% CI 0.4495 1.1994] (t = 4.56, p =.0002) with an estimated variance τ^2^ = 0.1709 and heterogeneity I^2^ = 77.9% (Q(22)=99.45, p <.0001). Looking only at the 18 studies comparing directly meditators to control (N=395 vs. 454), the effect size was slightly larger: g=1.025 [0.6055 1.4445] t=5.16 p<.0001, τ^2^ = .1053 I^2^ = 75.4% Q(17)=69.11, p <.0001 (figure 1).

**Figure 1.**
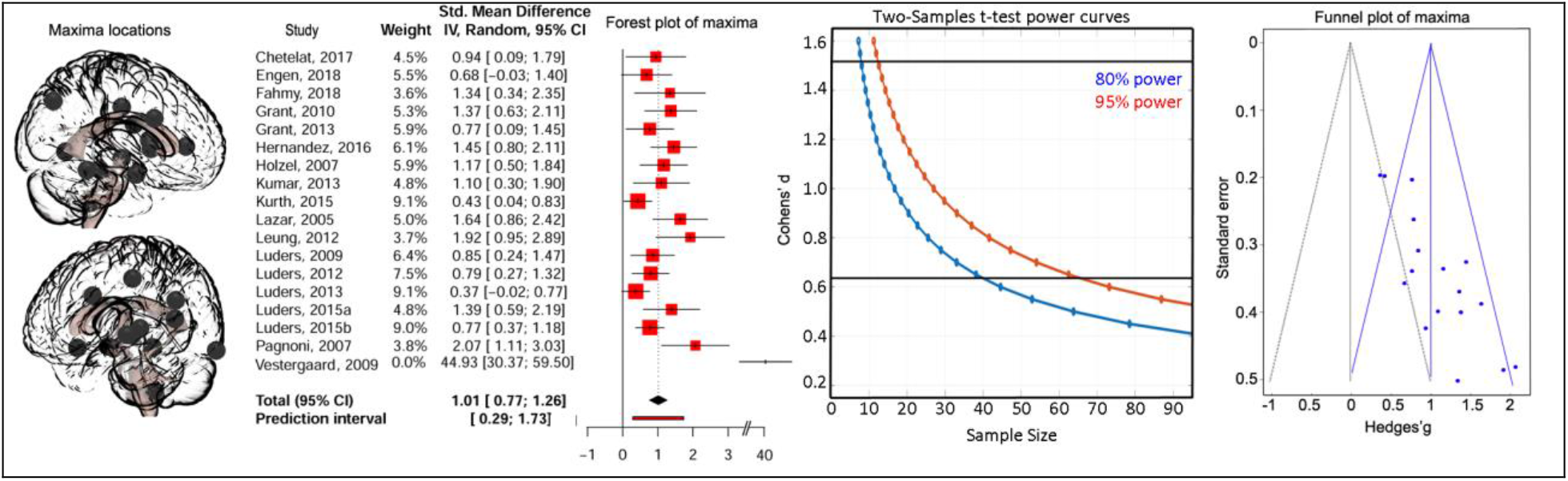
Whole brain maxima analyses. The left side of the figure shows location of maxima reported anywhere in the brain for studies comparing directly meditators to controls (n=18), leading to estimate an overall strength of evidence (forest plot of maxima – middle left) from which one can estimate how many subjects are needed in future studies (power curves with black horizontal bars representing the 95% CI bounds of the meta-analysis effect – middle right), knowing however that there is an publication bias since only significant results were used (funnel plot excluding Vestergaard-Poulsen et al., 2009 (weight = 0) with the null effect shown with the grey triangle vs. computed effect in blue – right hand side).

#### Selection bias

unrepresentative samples limit our ability to generalise results, which was apparent in most of the studies in this review. Examples include the use of healthcare workers as controls, samples consisting solely of volunteers or college students and geographically-limited recruitment. There are also instances where recruitment procedures are not described. A number of studies have included controls from databases of healthy adults, and while these may seem appropriate, there is concern over their prior experience with meditation given around 8% of the general (western societies) population meditation (Clarke et al., 2015). Since this is not specified, in some studies, it is not possible to assess whether or not the controls are appropriate (Martínez-Mesa et al., 2016). Another issue is the inclusion of participants from various or unspecified meditation backgrounds within the same study, which implies that various meditation/mindfulness disciplines are equal in effect -- as only acknowledged as a limitation in only one study (Chételat et al., 2017).

#### Information bias

is commonly seen with self-reporting, which may reduce study validity (Althubaiti, 2016). Most studies in this review recorded participants’ prior meditation experience in hours or years, this may be problematic because reliance on memory and honesty reduces objectivity, and increases the risk of including incorrect information. Studies using mindfulness questionnaires may also add to this problem, for example the five facet mindfulness questionnaire (FFMQ), which was shown to have issues with construct validity (Goldberg et al., 2016). Furthermore, questionable classification methods increase the risk of biased or incorrect information: for example, one study divides participants into high versus low frequency meditation/prayer groups based on no established classification method.

#### Attrition bias

Several cases of unaccounted dropouts have, again, limited the formation of a definite conclusion regarding meditation and GM. Although there are situations where dropouts may not affect bias (Bell et al., 2013), this review includes cases where bias is likely (see Table 2).

**Table 2.** Bias analysis showing for each of the 25 studies (✓ indicates bias, X indicates no bias, ? unclear).

|  | *Selection bias* | *Information bias* | *Attrition bias* | *Confirmation bias* |
| --- | --- | --- | --- | --- |
| **Chételat, 2017** | ✓ | ? Most likely that info was obtained by interviewing participants | X | ✓ |
| **Engen, 2018** | ✓ | ✓ | X | ✓? No mention of blinding |
| **Fahmy, 2018** | ✓ | ✓/MBI which was adapted from the original MBSR | ✓ | ? No mention of blinding, but also no mention of manual input |
| **Fayed, 2017** | ✓ | ✓ | X | X (check in text citations to match this) |
| **Friedel, 2015** | X | ✓ | ✓ (245 agreed, 82 final) | ✓ |
| **Grant, 2010** | ✓ | ✓ | X | X |
| **Grant, 2013** | ✓ | ✓ | X | X |
| Hernandez, 2016 | ✓ | ✓ | X | ? No mention of blinding during manual input |
| Holzel, 2007 | ✓ | ✓ | X | X |
| Kober, 2017 | ✓ | ✓ | X | X |
| Kumar, 2013 | ✓ | ✓ | X | ? Methods not clear-possible manual input. Blinding not mentioned. |
| Kurth, 2014 | ✓ | ✓ | X | X |
| Kurth, 2015 | ✓ | ✓ | X | X |
| Lazar, 2005 | ✓ | ✓ | X | ? Not mentioned |
| Leung, 2012 | ✓ | ✓ | X | ? Not mentioned |
| Lu, 2014 | ✓ | ✓ | ✓ scans with artifacts not included | ? Not mentioned |
| Luders, 2009 | ✓ | ✓ | X | X |
| Luders, 2012 | ✓ | ✓ | X | X |
| Luders, 2013 | ✓ | ✓ | X | ✓ likely. Manual input. Blinding Not mentioned |
| Luders, 2015a | ✓ | ✓ | X | Not mentioned |
| Luders, 2015b | ✓ | ✓ | X | ✓ likely. Manual input and no mention of blinding |
| Murakami, 2012 | ✓ | ✓ | X | X |
| Pagnoni, 2007 | ✓ | ✓ | ✓ image with artifact was not included | X |
| Taren, 2013 | ✓ | ✓ | ✓ 10 excluded due to missing data | ? semi-automated ROI segmentation. No mention of blinding. |
| Vestergaard-Poulsen, 2009 | ✓ | ✓ | X | X<br>No mention of manual input during segmentation. However the authors used histological samples. |

#### Blinding

The majority of studies were observational, retrospective and cross-sectional, where participant blinding was not performed since it was unnecessary. Of the studies that were longitudinal in nature, one did not mention participant blinding and the other did not involve any intervention, rendering this unnecessary. Investigator blinding was not required in studies using automatic methods of GM measurement, however those using semi-automatic measurement, or manual delineation of GM would require blinding since over/underestimation is possible with knowledge of the participant group. Some studies included here employed manual methods with no mention of observer blinding, which could lead to confirmation bias (Althubaiti, 2016).

#### Publication bias

as expected from analyzing maxima and illustrated on the funnel plot, the meta-analysis effect size is biased such as studies follow the expected null slope with weaker effects for higher precision studies and conversely stronger effects for lower precision studies. Importantly, no non-significant maxima were present because all studies, but one found effects, and the one not finding a difference did not report on effect size.

### Spatial meta-analysis and connectivity

Of the 25 studies included, 16 provided anatomical coordinates of regions showing significantly increased GM in meditators or following an intervention. Four studies had overlapping samples, with however no overlapping coordinates. The meta-analysis from all data demonstrated one significant cluster in the short insular gyrus extending to the claustrum (from [26 8 −18] to [38 14 −8] with a peak at [32 10 −14]). Overall, 6 studies reported effects in the right insula, but only 3 of them participated in the ALE cluster. All 6 studies were nevertheless neighbouring coordinates, all were used to create a summary functional connectivity map. This analysis showed significant association (i.e. above the 88% chance level) of the left insular cortex, the left/right inferior post-central gyri, the anterior cingulate gyrus and adjacent paracingulate gyri with the right insula (figure 2). Lowering the threshold to the ‘chance level’ 50% also show possible association with left and right frontal poles.

**Figure 2.**
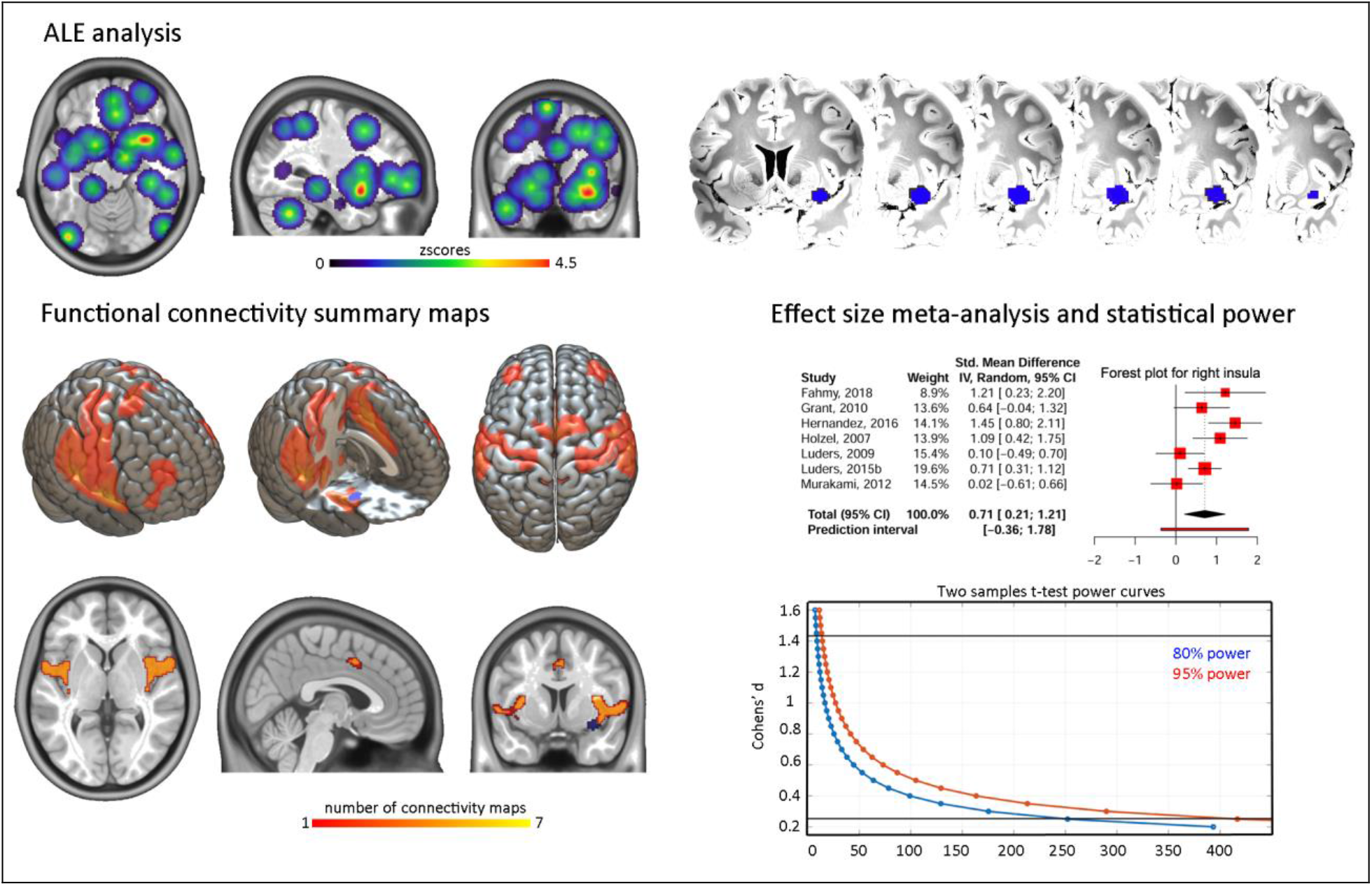
ALE analysis and derived results. The top of the figure shows the z-scores ALE for all coordinates entered into the analysis, leading to a significant result over the right short insular gyrus. From this region, coordinates from 7 studies (3 studies in the ALE results + 4 located posterior and dorsally) were used to obtain functional connectivity maps (bottom left) showing overlap with observed coordinates over the left insular and anterior cingulate/paracingulate cortices. The brain render shows the unthresholded summary map while slices display significant regions only. Effect sizes from these 7 studies were also entered into a meta-analysis (bottom right) to estimate sample size for future studies (power curves with black horizontal bars representing the 95% CI bounds of the meta-analysis effect transforming Hedges’g reported in the forest plot to Cohens’d).

## Discussion

Our meta-analysis indicates that meditation has a medium to large effect on grey matter (Cohen’s lower estimate ∼0.45 for all studies to higher estimate of ∼1.5 for studies comparing directly controls to meditators) with however a large heterogeneity in effect sizes (∼76%) and in location throughout the brain (figure 1). The only one area showing spatial consistency, i.e. the right anterior ventral insula (figure 2), have an effect size estimate between 0.2 and 1.4. Our results are concordant with both, an effect of meditation on expected brain areas, and a high spatial variance caused by methodological biases.

### Effect of meditation

The ALE meta-analysis reveals a significant cluster over the right anterior insula, going over the extreme capsule to the claustrum, an overlap from ROI from Fahmy et al. (2018 - opioid patients meditators > opioid patients control in the claustrum), Hernández et al. (2016 - mediators > controls in the short insula gyrus) and Murakami et al. (2012 - correlation with mindfulness in the short insula gyrus), a sub-region typically associated with emotion (Kurth et al., 2010). Lazar and colleagues (2005) were the first to observe important structural changes associated with sustained meditation practice in this region (although not included in the ALE analysis because no coordinates were reported in the article), but also the auditory, somato-sensorial and prefrontal cortices. The other studies reviewed reporting right insular differences (Grant et al., 2010, Hotzel et al., 2007, Luders et al., 2007, 2015) were more posterior or dorsal, sub-regions associated with sensori-motor afference or empathy (Kurth et al., 2010), altogether arguing for GM changes associated with classical cognitive effects of meditation (Srinivasan, 2019). While such insular changes can be expected from meditation experience, it is important to note that our meta-ALE-analysis is likely lacking statistical power with 16 studies included while estimates suggest ∼20 studies for reliable results (Eickhoff et al. 2016). However, the proximity of these 4 additional studies to the ALE cluster and the functional connectivity results (see below) lead to conclude to a true effect rather than a false positive. In this context, it is also interesting to note that cognitive traits associated to the right insula are also typically associated with the left insula, which we showed to be functionally connected with the right insula. Four studies in our review also reported left insular changes (Engen et al., 2018; Fahmy et al., 2018, Grant et al., 2010, Luders et al., 2015) but they were spatially too dispersed to show significant ALE overlap (figure2). Similarly, about 1/3 of studies (Chetelat et al. (2017), Engen et al., (2018), Fahmy et al., (2018), Grant et al., (2010), Grant et al., (2013), Kurth et al., (2014), Lu et al., (2014)) showed activations over the (broadly speaking) anterior cingulate/paracingulate cortices, without enough spatially consistency to be a significant region in the ALE analysis, but coordinates overlap with the functional connectivity maps. Given that this region has also been linked to meditation in other reviews (see Boccia et al., 2015) we therefore recommend to also considered it as a-priori region of interest in future studies.

### Spatial heterogeneity of effects

More often than not, the strongest effects were observed in different locations in the brain which may be partly attributed to a lack of statistical power. In our main effect size meta-analysis, only the strongest effects were used leading to a dispersion of effects (Hedges’ g from 0.18 to 2.07 - excluding Vestergaard-Poulsen et al., 2009) along the expected null line of the funnel plot. This indicates that the population effect size is overestimated, calling to use the lower bound of the confidence interval as a safe assumption on how many subjects are needed to properly estimate/quantify the effect of the influence of meditation on grey matter. For 95% statistical power, 62 subjects per group (124 in total) would be necessary, which is more participants than most studies have used. If one were to plan a study focusing on the insula, our analysis suggests up to 417 subjects per group (834 subjects in total using the lower bound of the CI). Because lack of power increases false positives (Button et al., 2013; Pernet, 2017) and because of the non-stationary nature of the grey matter values derived from the MRI signal (Hayasaka et al., 2004), larger effects than real ones (i.e. false positives) can be observed at random locations, explaining the observed spatial heterogeneity. Other factors related to design and analysis might also contribute to this variability (see below).

### Strengths and weaknesses of methods

This systematic review utilised multiple databases for the literature search and excluded studies that involved movement therapy as part of the meditation/mindfulness intervention. These factors increase the likelihood that all relevant studies were included and that the results obtained were not affected by confounding factors. On the other hand, it should be noted that of the studies included here, some involve participants with pain disorders and other illnesses, as well as adolescents, all of which could possibly bias results. Compared with the most recent ALE analysis (Boccia et al., 2015), we included more studies and used the updated software version that has a stricter type 1 Family-Wise error rate (permutation of maxima rather than FDR – which is the recommended setting, Eickhoff et al., 2016), and thus it would not be reasonable to expect too many similarities given those restrictive criteria.

### Moving forward research on the effect of mindfulness on the brain

The biggest concern on the observed results is the spatial variability across studies. As discussed above, the total sample size is an important contributing factor and future studies must power up if one wants, for instance, to demonstrate up or down regulation of a given brain region by meditation in patients. The bias analysis also revealed issues with (i) demographics, (ii) analyses, (iii) reporting, and (iv) experimental design. Variation in age, sex, ethnicity, education, socio-economic status, handedness and illnesses may confound the data, leading to the creation of inaccurate results. Apart from this, many studies did not include important demographic information, creating additional ambiguity surrounding the data and any conclusions that may be drawn from it. It is also important to consider the impact of neurological and psychiatric illnesses, as these may affect GM. The studies reviewed that did not report these were excluded, and of those that do mention exclusion, not all specify the measures used to assess such illnesses. Another important issue is the variability of meditation practices. It may be possible that some practices have more profound effects than others. Since only a limited number of studies with different meditation practices is available, it is not possible to assess whether this is true, or if any differences among practices exist at all. When studying meditation, we must acknowledge that experienced participants are likely to have had exposure to many practices over the years, and so it may be incorrect to assume that the practice measured at the time of the study is responsible for the differences seen. Furthermore, one must also keep in mind that accurate measurement of meditation itself is not entirely possible and concerns surrounding the validity of the various measurement tools used have been raised (Park et al., 2013). Because of the variability in demographics, it is also essential to include covariates in the brain data analyses. Less than half of studies however reported the inclusion of factors such as age, gender and total intracranial volume although those are essential to interpret results, no matter if significantly different between groups (Pernet, 2018). While a few articles made data available, most do not mention data sharing. Essential to reproductive and cumulative science (Nichols et al., 2017), details of the analyses are often missing (for instance having to figure out if and what covariates were used) and summary statistics maps never available (for instance sharing on NeuroVault (Gorgolewski et al., 2015) the unthresholded t-value map of difference controls vs. experts which would allow computing effect sizes for any brain region). Finally, in terms of design, the majority of studies that met the inclusion criteria were cross-sectional and observational. Although this is enough to show the presence of an effect, there is a need for more randomised controlled trials and longitudinal studies to demonstrate therapeutic benefits. Longitudinal studies should span lengthy periods of time, and involve regular scan intervals in order to examine effects over time. Despite their appeal, there are also a number of issues making them problematic. Such studies were performed over a condensed period of time, due to cost and time limitations, and an increased likelihood of drop-outs, making it difficult to determine long-term effects. At present, there is no defined interval time after which it is agreed that the effects of meditation should be apparent, and we have not yet concluded whether the effects of meditation practice are cumulative, that is, whether the reported changes are related to the amount of experience alone. Furthermore, we have not come to a conclusion on whether the effects are long-lasting beyond the intervention time frame. Improvements in study design, such as blinding are also essential in order to move forward.

## Conclusions

There is mounting evidence that meditation induces functional and structural changes in the brain. Our review shows that effects are relatively strong for structural changes ([0.45 1.5]) with consistent changes observed in the right insula, but methodological improvements are required to establish mindfulness meditation as a therapeutic tool.

## Data Availability

All of the data necessary to replicate what is presented here is available (extracted data, R scripts, functional connectivity maps and Matlab code).

https://github.com/CPernet/meta_meditation

## Availability of data and materials

All of the data necessary to replicate what is presented here is available @https://github.com/CPernet/meta_meditation (extracted data, R scripts, functional connectivity maps and Matlab code).

## Acknowledgments

Thank you to D. Lakens for publishing YouTube tutorial videos on meta-analysis and bias.

## Author Contributions

CP and AZ conceptualized the review and conducted the ALE analysis, AZ conducted the systematic review and bias analysis, CP and NB conducted the statistical meta-analyses and AD helped edit the manuscript. All co-authors have contributed to the preparation of the manuscript, provided important intellectual content and read and approved the final version.

